# Effectiveness and clinical relevance of kinesio taping in musculoskeletal disorders: A protocol for an overview of systematic reviews and evidence mapping

**DOI:** 10.1101/2024.03.07.24303944

**Authors:** Qingcong Mo, Siqi Xu, Fangfei Hu, Xiaoyan Zheng

## Abstract

**Introduction:** Kinesio taping (KT) has been extensively applied in the management of musculoskeletal disorders. Although plentiful systematic reviews (SRs) have evaluated its efficacy, there are no convincing conclusions due to disperse and inconclusive results, and its clinical relevance remains unclear. Hence, there is a need to summarise all the SRs for the comprehensive and consistent evidence. This overview aims to appraise the overall effectiveness of KT in musculoskeletal disorders, and provide evidence maps to visualise the findings.

**Method and analysis:** Electronic databases (Cochrane Database of Systematic Reviews, MEDLINE, Embase, Epistemonikos, PEDro, Scopus, ISI Web of Science) and reference lists will be searched from inception to September 2024 for the SRs of randomised controlled trials (RCTs). The SRs involving comparisons of the effectiveness between single or adjunctive KT and other interventions for patients with musculoskeletal disorders will be included. The primary and additional outcome to be considered will be the core outcome set, and the patient-reported outcome measure and patient-important outcome, respectively. Two reviewers will independently screen and select studies, extract the data, and evaluate the reporting and methodological quality of eligible SRs as well as the risk of bias of included RCTs. For the SRs without meta-analysis, we will collate the number of RCTs that showed any differences in outcomes. For the SRs with meta-analysis, we will provide the original summary of evidence (e.g., pooled effects and heterogeneity) for outcomes with an evaluation of missing results and clinical relevance. The certainty of each outcome will be measured, and user-friendly maps of findings will be presented graphically.

**Ethics and dissemination:** Formal ethical approval for this study is not required, since the data will be only collected from published literature in public databases. The results will be disseminated in the peer-reviewed academic journal, and relevant datasets will be preserved in the online repository.

**PROSPERO registration number:** CRD42024517528

**Strengths and limitations of this study:**

● This study will be carried out and reported following the Cochrane Collaboration Handbook for Overviews of Reviews and the Preferred Reporting items for Overviews of Reviews statement.
● This study will only include systematic reviews with and without meta-analysis of randomised controlled trials.
● This study will evaluate the reporting and methodological quality of systematic reviews, examine the risk of bias and degree of overlap of randomised controlled trials, and manage discordant results by selecting the most representative research.
● This study will perform the assessment of missing results and certainty for the evidence, interpret the clinical relevance between effects and outcomes, and provide evidence maps of comprehensive and systematic findings.
● This study may not be up-to-date as newly published randomised controlled trials will not be included, and ultimate conclusions will probably be limited by clinical heterogeneity due to variations in dose or application method of kinesio taping.

## INTRODUCTION

Musculoskeletal disorders are a major and serious healthcare issue.[1] It consist of a group of more than 150 different and irreversible diseases/conditions that affect the locomotor system (including muscles, bones, joints and adjacent connective tissues).[2,3] Among them, the most prevalent conditions are low back pain, fractures, osteoarthritis, other injuries, neck pain, amputation, and rheumatoid arthritis.[4,5] Globally, musculoskeletal disorders are common among adolescents to the elderly, and the highest burden is expected to be concentrated in people aged 50-54 years.[6,7] Over the past few decades, with the growth and aging of population, musculoskeletal disorders have become increasingly serious.[4,5]

Estimates of the prevalence of musculoskeletal disorders suggested substantial increases in global cases and years of life lived with disability, affecting approximately 1.71 billion individuals.[4,5] Patients with musculoskeletal disorders may suffer from various health-related negative outcomes, involving problems in pain intensity, functional status, psychological state, quality of life, and more.[5,8,9] The consequences of these problems may lead to work-related disability while reducing productivity and increasing direct and indirect economic costs.[3,8,10,11] In addition, the risk of developing other noncommunicable diseases, especially cardiovascular diseases, is higher in people with musculoskeletal disorders than in those without them.[12] As a typical feature of musculoskeletal disorders, co-occurring persistent pain in different body regions beyond the primary pain may have implications for patients’ prognosis, treatment method, and outcome.[13,14] Hence, research on musculoskeletal disorders requires more attention, although they mainly lead to disability rather than death.[15,16]

In an attempt to meet the massive demand of rehabilitation services,[4,5] active management for musculoskeletal disorders is needed.[16–18] Currently, non-invasive treatments have a significant impact on the treatment of various musculoskeletal disorders.[15] Among them, kinesio taping (KT), a form of taping technique, has been extensively utilized across countries/regions for many years to support sports science and rehabilitation.[19] The application of KT in musculoskeletal disorders is widespread.[20]

### Description of the intervention

KT is an elastic therapeutic taping tool consisting mainly of cotton, originally created and introduced by Dr. Kenso Kase in the 1970s.[21] It is lightweight, waterproof, breathable and available in a wide range of colours, types, lengths, widths, textures and techniques.[21] Following assessment and instruction from healthcare professionals, KT can be directly fixed to the target tissue with different combinations of cuts (e.g., Y, I, X tape), tensions (e.g., percentage of stretch), and directions (e.g., muscle insertion to origin).[21] Wearing of KT may maintain the effect for 3 to 5 days, ideally following the 24 hours application rule based on the skin condition.[22]

Over the past decade, the popularity of KT is increasing worldwide given the promising benefits suggested by plentiful clinical studies.[19,22] KT assists users with musculoskeletal disorders in relieving pain intensity,[23] modulating extremity function,[23] and improving proprioception,[24] stability,[25] range of motion,[26] and quality of life.[27] These therapeutic effects are probably supported by the mechanisms of KT that it increases the subcutaneous space, stimulates skin sensory receptors, and provides support for mechanical behaviour and biomechanics of the skin.[21,28–30] Applying different tensions of it on the skin may reduce partial pressure, accelerate blood and lymphatic circulation, and increase muscle temperature, thus promoting regional microcirculation.[21,31,32] People with a psychologically anticipated response to the effect of KT are likely to induce greater placebo effects, thus contributing to enhanced muscle function.[33–35] Although there is some evidence to support the theory of KT, its precise working mechanism remains unclear and many studies showed conflicting therapeutic results.[36] Consequently, the inconclusive findings of KT have resulted in a longstanding debate regarding the effectiveness and clinical relevance.

### Significance of conducting the review

Due to substantial controversy over the results of KT reported in the primary studies, researchers have carried out plentiful systematic reviews (SRs) with and without meta-analysis to evaluate the clinical effectiveness of KT.[37–40] However, the inconclusive evidence of musculoskeletal disorders was presented in the SRs as well, involving elbow,[41,42] shoulder,[43–45] back,[46–48] knee,[49–51] and spine.[52]

Meanwhile, the majority of SRs may not examine the clinical relevance of KT. The minimal clinically important difference (MCID) is considered to be the smallest change that appears to have a benefit implication for patient’s treatment outcome.[53,54] The application of MCID assists in establishing numerical thresholds for clinical research that only identified statistically significant impacts of intervention with little or unknown clinical relevance for patients.[55–57] Nevertheless, Embry and Piccirillo retrieved the journal and found that large percentage of randomised controlled trials (RCTs) (69%) did not define the MCID or mention it.[58] The lack of additional confirmation regarding the clinical relevance of KT presumably added difficulties to clinical interpretation of its effects.

As the number of SRs increased to date, the existing evidence of KT probably created gaps between research and practice, as well as variations in clinical opinions among healthcare providers.[59,60] Hence, there is a need to summarise and assess the overall effectiveness of KT in musculoskeletal disorders from all the SRs.

Overview of SRs, a method of reviewing multiple SRs, is capable of collating broad information, synthesizing diverse findings, and presenting a well-organized and detailed summary of evidence.[61,62] This evidence-based method is frequently applied to develop a unified perspective when there are conflicting conclusions on a specific topic at the level of the SRs due to different selection methods, potential sources of heterogeneity and risk of bias, and variable reporting and methodological quality.[61,62] In addition, evidence mapping is a effective and evolving methodological tool for integrating and presenting evidence through user-friendly visual graphics.[63,64] Cupler and colleagues have provided an evidence map of four types of taping (KT, Rigid taping, McConnell taping and Mulligan taping) for musculoskeletal conditions.[65] However, the limitation of the study is that the evidence maps were based on the included RCTs and did not clarify the relationship between the outcomes and the contradictory findings of the SRs. Application of evidence mapping for overview of SRs plays a complementary role in conveniently identifying research gaps and rapidly disseminating knowledge.[63,64,66,67]

To date, no studies have evaluated the existing evidence on the effectiveness and clinical relevance of KT in musculoskeletal disorders from the SRs. To fill the gap of knowledge, we will conduct an overview of SRs and evidence mapping to inform evidence-based clinical practice and support healthcare decision-making. The relevant findings obtained by this study will be expected to benefit researchers, physiotherapists, stakeholders, and particularly multiple patients at various stages of musculoskeletal rehabilitation.

### Objective and research question

This study aims to appraise the overall effectiveness of KT in musculoskeletal disorders, and provide evidence maps to visualise the findings. This study will examine the following research questions: (1) Is KT effective in patients diagnosed with musculoskeletal disorders? (2) What types of outcomes does KT demonstrate positive, no, or negative effects on?

## METHODS AND ANALYSIS

### Protocol and registration

This study is designed as an overview of SRs and evidence mapping. We have prospectively registered this protocol within the International Prospective Register of Systematic Reviews (PROSPERO, Registration number: CRD42024517528). The protocol has been presented in accordance with the Preferred Reporting Items for Systematic Reviews and Meta-Analysis Protocols (PRISMA-P) statement (Supplementary File 1).[68] We will conduct the overview following the methodology of Cochrane Collaboration Handbook for Overviews of Reviews.[69]

### Patient and public involvement

No patients or the public involved in this overview of SRs and evidence mapping.

### Eligibility criteria

#### Types of studies

Standard-compliant SRs with and without meta-analysis of RCTs will be considered for inclusion. The definition of systematic review and meta-analysis will be adhered to the Preferred Reporting Items for Systematic Reviews and Meta-Analysis (PRISMA) 2020 statement,[70] as follows: systematic review means to use explicit, systematic methods to collate and synthesise findings of studies that address a articulated question; meta-analysis is a statistical method for synthesising results when research effect estimates and their variances are available. If the SRs were published in other languages than English, or only searched one database, or no details of primary RCTs were provided, they will be excluded. Protocols of overview and SRs, scoping review and network meta-analysis will be excluded as well.

#### Types of participants

Guided by the International Classification of Diseases 11th revision (ICD-11),[71] we will include participants who have a diagnosis of musculoskeletal disorders. Eligible individuals will be included, regardless of their age, gender, and region. Additionally, we will exclude animals from participation.

#### Types of interventions

The SRs that evaluated the effectiveness of KT in musculoskeletal disorders will be considered for inclusion. KT must be used as a core treatment method in the experimental group, either as monotherapy or combination therapy. The way of applying KT on the body must be related to the musculoskeletal system. It will not be limited by tension, direction, and regimen.

#### Types of comparators

No strict intervention limitations on the type of control group will be set, such as no treatment, standard of care, placebo control, medication therapy, rehabilitation, and other interventions (e.g., traditional Chinese medicine therapy, surgical treatment). KT as a treatment method will be excluded in this group.

#### Types of outcome measures

With the ability to lower the risk of heterogeneity, inconsistency, and outcome-reporting bias between trials, the core outcome set will be the primary outcome.[72] First, we will search specific musculoskeletal disorders related to KT usage in eligible SRs on the MEDLINE or the website (https://comet-initiative.org/). Then, we will select the most up-to-date (i.e., date approaching 2022-2024), credible (e.g., registered research, multi-round survey, more stakeholders included), and standard (e.g., reported more complete items in the checklist of Core Outcome Set-STAndards for Development)[73] core outcome set to consider any promising outcomes that must be relevant to patients.

If partial musculoskeletal disorders are not assessed by the core outcome set, we plan to consider patient-reported outcome measures [74] and patient-important outcomes.[75] They are able to reflect patient perspectives on their symptoms, functional status, quality of life, and more. The following additional outcomes will be considered:

1. Pain intensity, which measured by standard or specific scales (e.g., Numerical Rating Scale (NRS), Visual Analogue Scale (VAS)).
2. Upper and lower limb function and/or disability status, which measured by standard or specific scales (e.g., Range of Motion (ROM), Disabilities of the Arm, Shoulder and Hand (DASH), Victorian Institute of Sport Assessment-Patella Questionnaire (VISA-P), Roland-Morris Disability Questionnaire (RMDQ)).
3. Quality of life, which measured by standard or specific scales (e.g., Short-Form 36 Health Survey Questionnaire (SF-36), EORTC Core Quality of Life questionnaire (EORTC QLQ-C30), WHO Quality of Life Brief Questionnaire (WHOQOL-BREF)).
4. Disease-specific symptom, which measured by standard or specific scales (e.g., Grip Strength (GS) for lateral epicondylitis).

### Search strategy and selection

According to the Preferred Reporting Items for Systematic reviews and Meta-Analyses literature search extension (PRISMA-S) guideline,[76] our search will be conducted in the following electronic databases (Cochrane Database of Systematic Reviews, MEDLINE, Embase, Epistemonikos, PEDro, Scopus, ISI Web of Science) from inception to September 2024. We will also perform a search to identify any registered yet unpublished SRs and grey literature in the PROSPERO and OpenGrey, respectively. In addition, we will hand-search the reference lists of included SRs for any relevant cited SRs in Google Scholar. The search strategy is developed involving the integration of MeSH terms, keywords and free text terms related to KT: (kinesio taping OR kinesio tape OR kinesiotaping OR kinesiotap OR physiotape OR tape OR taping) AND (systematic review OR meta-analysis).

Two independent literature reviewers will screen titles and abstracts of the SRs output through the EndNote V.21 software to identify eligible articles. Then, the same reviewers will download the promising SRs and assessed them by full-text reading for final inclusion. Afterwards, applying Gwet’s AC1 statistics and related 95% confidence interval (CI),[77] one reviewer will evaluate the agreement on study selection between two reviewers using R language V.4.3.2. Considering that application of the classification from Landis and Koch to Gwet’s AC1 is inappropriate,[78] the inter-rater reliability will be cautiously explained. The Fisher’s exact test will be performed to determine its statistical significance (P value). Any disagreement will be discussed or consulted the third reviewer for a consensus.

### Data extraction

We will extract and cross-check data from the SRs in standardised tables based on predefined criteria, using an electronic form of Microsoft Excel 2019. All data must be related to KT only. Data concerning the basic information (first author’s name, year of publication, country/region of the first author’s affiliation, and number of the included primary RCTs), participants characteristic (number of participants and disease or condition), search strategy (number and names of databases searched and date range of search), type of comparison, outcome measurement, evidence assessment tool (risk of bias and certainty of evidence) and study conclusion (e.g., positive effect, no effect, negative effect) will be extracted. If we identified discrepant data of included same studies in different SRs, we plan to contact the corresponding authors or to retrieve and follow the raw data of primary RCTs.

If the SRs with meta-analysis analysed outcomes with more than one study, two reviewers will extract the analytical methods (fixed or random effects model), heterogeneity (Cochran’s Q test P value and I-square statistic), pooled effects with 95% CI (mean difference (MD) or standardized mean difference (SMD) for continuous outcome; relative risk (RR) or odds ratio (OR) for binary outcome), direction (differences in favour of KT or control intervention), statistical significance (P value) and publication bias (at least 10 studies). If the follow-up data is available, we will extract them as well.

To provide a complete summary of KT, we will use the Risk of Bias due to Missing Evidence (ROB-ME) tool to assess the meta-analysis results.[79] Briefly, three steps will be conducted following the tool: select meta-analyses related to outcome (step 1), determine which eligible studies have missing results (step 2), and consider potential for missing studies (step 3). Response options for the signal questions are categorised as ‘yes’, ‘probably yes’, ‘probably no’, ‘no’, ‘no information’ or ‘not applicable’. Then, the results will be judged and interpreted as ‘low risk of bias’, ‘some concerns’ or ‘high risk of bias’. Discrepancies considered potentially relevant by one reviewer will be discussed and, if needed, will be ultimately resolved by the third reviewer.

### Managing the overlap between primary RCTs

As the number of SRs and updated SRs for KT increased to date, the identical or highly similar questions related to musculoskeletal disorders were probably answered.[80] To avoid double counting data from overlapped SRs, we will extract the information on characteristics of primary RCTs separately (first author’s name, year of publication, number of participants, type of comparison, pre– and post-intervention outcome measurement data with or without follow-up). After removing duplicated articles, we will combine the results into a table.

Based on the specific clinical question, a matrix of included primary RCTs in the SRs will be created and visualised to evaluate the amount of overlapping using the Graphical Representation of Overlap for OVErviews (GROOVE) tool.[81] When collating each outcome in the matrix, we will calculate the corrected covered area (CCA) regardless of any structural missing data. The CCA refers to a measurement for the degree of overlap and it will be interpreted as ‘slight overlap’ (0%–5%), ‘moderate overlap’ (6%–10%), ‘high overlap’ (11%– 15%) or ‘very high overlap’ (>15%).[82]

### Assessment of the reporting quality

Transparent and complete reporting of SRs supports to examine the feasibility of the methods and the reliability of the findings, thus bolstering evidence-based decision-making.[70,83] Two reviewers will separately evaluate the reporting quality of SRs with and without meta-analysis in accordance with the Synthesis without meta-analysis (SWiM) reporting guideline [83] and the PRISMA 2020 statement.[70] The SwiM guideline consists of 9 items and contains critical features of synthesis in the SRs without meta-analysis (e.g., the method of study category, data presentation, finding summary). The PRISMA checklist covers 27 items and is designed for SRs with meta-analysis that require detailed reporting in different sections (e.g., abstract, methods, results). Each item of the above checklist will be recorded and graded as ‘completely reported’, ‘partially reported’ or ‘not reported’ depending on whether the domains were clearly documented. Then, we will calculate the compliance rate of the reporting in each of SRs to overview the overall reporting quality. Any differences of opinion will be resolved through discussion or determined by the third reviewer.

### Assessment of the methodological quality and the risk of bias

The quality assessment of eligible SRs includes both the methodological quality and the risk of bias [84]. Two reviewers will independently assess them by the A Measurement Tool to Assess Systematic Reviews 2 (AMSTAR 2) [85] and Risk of Bias in Systematic reviews (ROBIS),[86] respectively. The AMSTAR 2 is a valid, reliable and user-friendly appraisal tool and most of the questions are binary.[87] After answering 16 original items (recommended critical domains are 2, 4, 7, 9, 11, 13, and 15), the overall methodological confidence of SRs will be judged as follows: ‘high’ (no or one non-critical weakness), ‘moderate’ (more than one non-critical weakness), ‘low’ (one critical flaw with or without non-critical weaknesses) or ‘critically low’ (more than one critical flaw with or without non-critical weaknesses). The ROBIS has rigorous methodology and is intended for assessing the level of bias within SRs through three phases: assess the relevance of SRs to outcome for KT (phase 1), identify concerns with the review process including study eligibility criteria, identification and selection of studies, data collection and study appraisal, synthesis and findings (phase 2), judge the risk of bias based on the concerns of each domain bias (phase 3).[86] The overall risk of bias will be rated as ‘low’, ‘unclear’ or ‘high’ according to the answers to signalling questions.

In addition, we plan to re-evaluate the quality of each primary RCTs using the Cochrane Risk of Bias assessment tool 2.0 (ROB 2.0),[88] if the SRs appraised the risk of bias by means of other tool (e.g., Cochrane risk of bias tool, Jadad scale, PEDro scale). The ROB 2.0 consists of five bias domain originating from the randomisation process, deviations from intended interventions, missing outcome data, measurement of the outcome and selection of the reported result. Each domain will be required to rank the levels of evidence bias through a series of signalling questions to determine the judgement of overall risk of bias as ‘low risk of bias’, ‘some concerns’ or ‘high risk of bias’.[88] Any differences will be settled through discussion or with the assistance of the third reviewer.

### Strategies for data analysis

Within the SRs without meta-analysis, we will collate and summarise the number of RCTs that demonstrated any differences in the outcomes (i.e., statistically significant positive effect [direction of effect in favour of KT], statistically significant negative effect [direction of effect in favour of control intervention] and non-statistically significant effect). Statistical significance will be defined as a P value less than 0.05.[89] If the SRs incorporated comparisons that did not meet the eligible inclusion criteria, the corresponding RCTs will not be applicable for calculation. Using the vote counting method, we plan to present these RCTs as a percentage of all primary studies in the SRs along with 95% CI (Wilson interval).[90]

Regarding the SRs with meta-analysis, we will present the original summary of evidence (e.g., pooled effects, heterogeneity and direction) without carrying new meta-analyses for outcomes. However, when the same conceptual outcome that included interventions beyond KT (e.g., Rigid taping, McConnell taping, Mulligan taping) or was reported by different measurements (e.g., outcome of interest that should be calculated in MD but was calculated in SMD), we plan to back-translate the primary data in the SRs to re-estimate the overall effects. For re-analysing the outcome data that incorporated other distinct types of KT, we will exclude these RCTs by examining the characteristics of primary studies. Then, the associated findings (i.e., pooled effects, heterogeneity and direction) will be extracted, and the publication bias (at least 10 RCTs) will be evaluated by the Egger’s regression test. For the conversion of appropriate outcome measurements, we will present the binary outcomes as RR since the usage of OR in RCTs may exaggerate the effect size, leading to the misinterpretation in decision-making.[91,92] And the continuous outcomes will be presented as MD (same conceptual outcomes are measured on same scales) or SMD (same conceptual outcomes are measured on different scales).[93] In the above procedures, to maximise the retention of the authors’ data processing methods, the re-estimation will be performed under the same settings corresponding to the SRs using the Review Manager V.5.4 software.

### Strategies for managing concordant or discordant results

To provide the concordant evidence, we will organise the final results of KT (i.e., the proportion of RCTs that showed any effect and the overall pooled effects) from the SRs with and without meta-analysis into three categories (positive effect, no effect, and negative effect).[69] Table 1 provides detailed criteria for classification. A threshold of 80% SRs in the same classification will be set to determine the concordance of results [94] and the robustness of review findings will be discussed.[95]

**Table 1.** Effects classification criteria. CI, confidence interval; RCTs, randomised controlled trials; KT, kinesio taping.

| Type of systematic review | Category of effect | Description |
| --- | --- | --- |
| Systematic review without meta-analysis | Positive effect | The proportion of RCTs that showed statistically significant effect in favour of KT is the highest |
|  | No effect | The proportion of RCTs that showed non-statistically significant effect is the highest |
|  | Negative effect | The proportion of RCTs that showed statistically significant effect in favour of control intervention is the highest |
| Systematic review with meta-analysis | Positive effect | Continuous outcome: the statistically significant pooled effect size and associated 95% CI is higher than 0 points (direction of effect in favour of KT) |
|  |  | Dichotomous outcome: the statistically significant pooled effect size and associated 95% CI is higher than 1 points (direction of effect in favour of KT) |
|  | No effect | Continuous outcome: the associated 95% CI of pooled effect size crosses 0 points |
|  |  | Dichotomous outcome: the associated 95% CI of pooled effect size crosses 1 points |
|  | Negative effect | Continuous outcome: the statistically significant pooled effect size and associated 95% CI is lower than 0 points (direction of effect in favour of control intervention) |
|  |  | Dichotomous outcome: the statistically significant pooled effect size and associated 95% CI is lower than 1 points (direction of effect in favour of control intervention) |

When there are discordant results, methods of treatment involves examining and recording discordance, using adjunct decision rules or tools, and selecting the most representative study among the SRs.[96] In this management, we will consider varied standards for choosing the best SR, as the new meta-analyses will not be conducted and the utilization of Jadad algorithm will be difficult to follow (i.e., limitations in operationalization and interpretation).[97] First, we will examine the resemblance of clinical question (e.g., moderate to very high overlap presented in CCA, similar scale used to combine effect size) across the SRs. Afterwards, the four-domain strategies (comprehensiveness, timeliness, riskiness, and reporting) will be applied to select the best SR that reported relatively trustworthy findings, as below:

1. Comprehensiveness: the SRs included the highest number of RCTs and participants.
2. Timeliness: the SRs conducted the most recent search date (i.e., date approaching 2022-2024) in the largest number of databases (e.g., MEDLINE, Embase, CINAHL).
3. Riskiness: the SRs that were evaluated as the highest methodological quality and the lowest risk of bias according to the AMSTAR 2 and ROBIS tool.
4. Reporting: the SRs reported the most complete items (number of items completely reported and partially reported) in the checklist of the SwiM guideline and the PRISMA 2020 statement.

### Assessment of the certainty of evidence

Certainty of evidence supports the clinical and health decision-making process and its assessment is an essential part of overview of SRs according to the Cochrane guidelines.[98,99] For the SRs without meta-analysis, we will assess the overall quality of evidence following the Grading of Recommendations, Assessment, Development, and Evaluation (GRADE) framework.[100] Details of the application of the GRADE framework in the SRs without meta-analysis are in Supplementary File 2.

If the SRs with meta-analysis used the GRADE methodology to determine the quality of evidence, we will extract and present the following information associated with outcomes: five domains of downgrade (risk of bias, inconsistency, indirectness, imprecision and reporting bias), three domains of upgrade (large effects, dose response and opposing plausible residual bias and confounding) and the corresponding certainty (high quality, moderate quality, low quality and very low quality).[101] In contrast, if these SRs did not report the certainty of evidence, we will evaluate the overall quality of summary of evidence by the GRADE methodology.

### Assessment and interpretation of clinical relevance

We propose comparing the continuous pooled effects with 95% CI from each meta-analysis to the MCID. With the increasing publication of MCID measurements involving different outcomes in musculoskeletal disorders,[102–105] there may be overlap. Hence, we will search and select the MCID that most closely resemble the outcomes and participants in electronic databases corresponding to our search strategy (e.g., MEDLINE, Embase, ISI Web of Science). Priority consideration will be given to the MCID determined through anchor-based methods. The MCID described as percentage change from baseline for outcomes will not be considered since this method may not be suitable for statistical analysis.[106] If the included MCID is appropriate, the pooled effects reported in MD will be directly compared to it. The pooled effects reported in SMD will not be compared to the MCID as various lumped scales may lead to substantial uncertainty. Nevertheless, when the pooled effects reported in SMD is greater than 0.5 (medium effect size), we will consider the difference between KT and other interventions as the MCID.[55,93] If the appropriate MCID is not available, we will select and use the standard deviation for distribution-based calculation.[55,56] Details of the calculation of clinical relevance are in Supplementary File 3.

Subsequently, we will adopt the systematic method provided from Man-Son Hing et al. to determine and interpret the clinical relevance of results (Figure 1).[107] The approach focuses on four levels of clinical importance between the MCID and the pooled effects with 95% CI, as below:

1. Definite: the lower limit of the 95% CI is greater than the MCID.
2. Probable: the lower limit of the 95% CI is smaller than the MCID, but the pooled effect size is greater than the MCID.
3. Possible: the pooled effect size is smaller than the MCID, but the upper limit of the 95% CI is greater than the MCID.
4. Definitely not: the upper limit of the 95% CI is smaller than the MCID.

**Figure 1.**
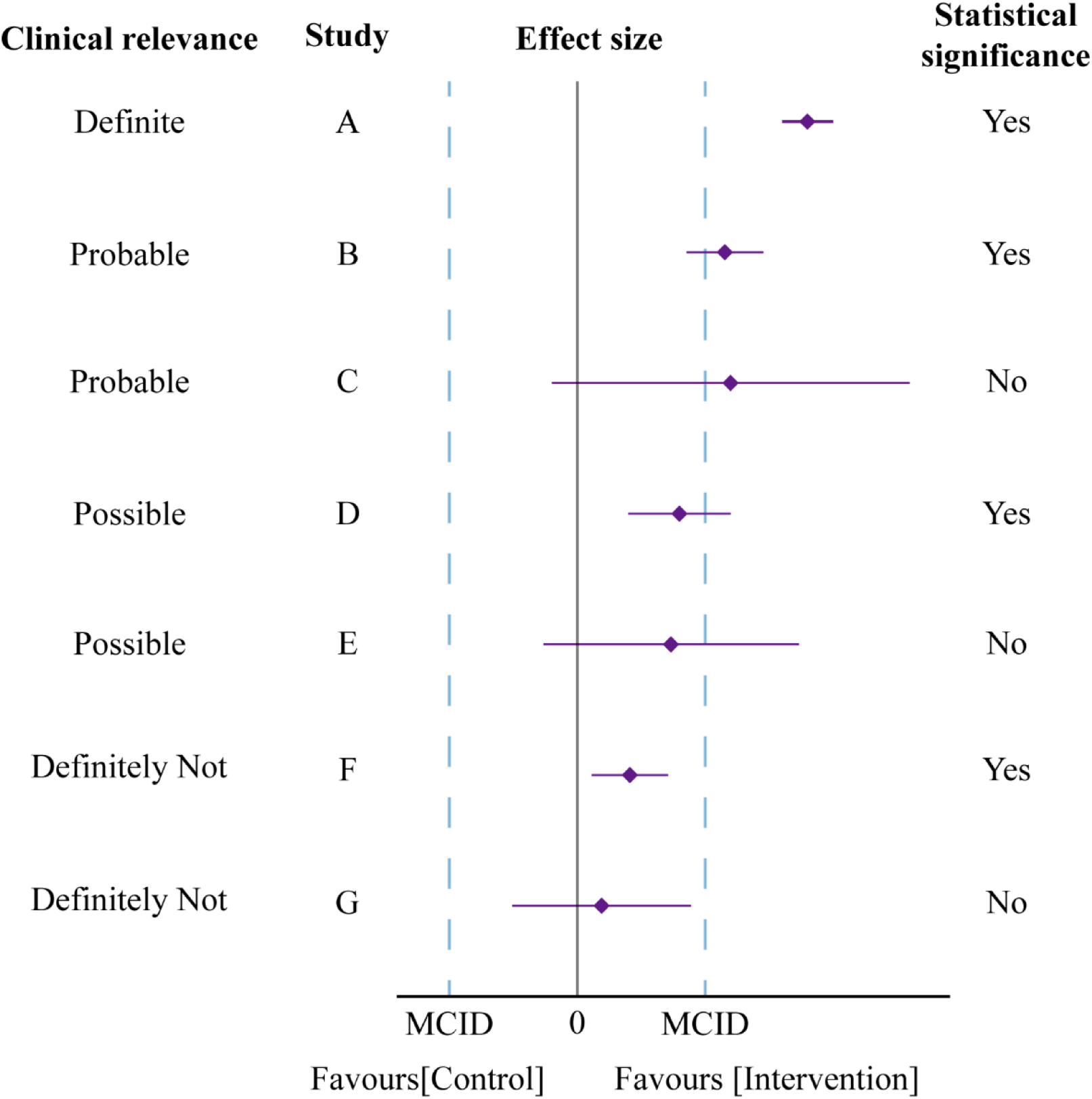
Method of interpreting clinical relevance developed by Man-Son-Hing et al.[107] and adapted. Dotted lines represent the clinical relevance threshold. MCID, minimal clinically important difference.

### Evidence mapping of findings

To provide credible evidence maps related to KT, we will summarise the evidence from the SRs and create user-friendly matrices of the relationship between effectiveness (positive effect, no effect, and negative effect) and outcomes. Then, we will grade the strength of evidence into five levels (convincing, highly suggestive, suggestive, weak, and non-significant evidence).[108–110] For the SRs without meta-analysis, we will evaluate the strength of evidence of positive or negative effects using the binomial test. This test (null hypothesis is equal to 0.5) will be applied to detect whether KT has a true effect.[90] For the SRs with meta-analysis, the strength of evidence will be assessed through strict criteria (e.g., number of participants and statistical significance). We will calculate the 95% prediction interval from the primary study specific data that corresponds to the SRs for outcomes. We plan to examine the excess of statistically significant bias by detecting whether the number of observed nominally significant studies differs from the expected number of research with significant results.[111] Details of the classification criteria for strength of evidence are in Table 2.

**Table 2.** Classification criteria of strength of evidence.

| Type of systematic review | Strength of evidence | Description |
| --- | --- | --- |
| Systematic reviews with meta-analysis | Class I: convincing evidence | Number of included participants>1000; statistical significance at $P<10^{-6}$; no large heterogeneity ($I^2<50\%$); 95% prediction interval not including null value; no small-study effects; no excess significance bias |
| | Class II: highly suggestive evidence | Number of included participants>1000; statistical significance at $P<10^{-6}$; largest study significant |
| | Class III: suggestive evidence | Number of included participants>1000; statistical significance at $P<10^{-3}$ |
| | Class IV: weak evidence | Statistical significance at $P<0.05$ |
| | Non-significant evidence | Statistical significance at $P>0.05$ |
| Systematic reviews without meta-analysis | Class I: convincing evidence | Probability of observing the evidence if positive or negative effects are not true at $P<0.05$ |
| | Class IV: weak evidence | Probability of observing the evidence if positive or negative effects are not true at $P>0.05$ |
|  | Non-significant evidence | Evidence of no effect observed |

### Strategies for evidence synthesis

We will report the overview following the Preferred Reporting items for Overviews of Reviews (PRIOR) statement.[112] The selection of the SRs will be reported using the PRISMA flow chart in accordance with the PRISMA-S guideline, and the associated inter-rater reliability (Gwet’s AC1 statistics) will be presented in narrative form. A list of included and excluded SRs will be provided with descriptive explanation of reasons.

The characteristics of the included SRs regarding the relevant details of basic information, participant characteristic, search strategy, type of comparison, outcome measurement, evidence assessment tool and study conclusion will be summarised and reported in tabular form. The matrix of evidence table at the outcome level will be visually presented in the heat map graphics that informed the degree of overlap between primary RCTs in the SRs.

We will report the results concerning the quality of the RCTs (risk of bias assessed by the ROB 2.0 tool) and the quality of the SRs (reporting assessed by the SWiM guideline and the PRISMA 2020 statement, methodology assessed by the AMSTAR 2, risk of bias assessed by the ROBIS tool) in tables. The final summary of evidence (the proportion of RCTs that showed any effect and the overall pooled effects) will be tabulated, along with the discussion of sensitivity to concordant or discordant results.

The effectiveness (positive, unclear, negative effect) of KT for each outcome of interest in musculoskeletal disorders will be presented accompanied by the clinical relevance and certainty of evidence using tables and figures. The visual evidence maps of evidence (the relationship between effectiveness and outcomes) with the corresponding strengths will be plotted, sub-grouped by different body regions (lower extremity, upper extremity, spinal conditions, and not otherwise classified).[65]

## ETHICS AND DISSEMINATION

This overview of SRs and evidence mapping does not require ethical approval, as the data will be only collected from published literature in public databases. This results will be published in the peer-reviewed academic journal, and relevant datasets will be preserved in the online repository.

## AUTHORS’ CONTRIBUTIONS

QM and SX, as co-first authors, made equal contributions to the study. QM, SX, FH and XZ contributed to the conception and design of the study. SX and FH designed the eligibility criteria and search strategies. QM, SX and FH developed the study quality evaluation, data extraction and analysis plan, and evidence presenting method. XZ is the study guarantor. All authors drafted the protocol and approved the final version for publication.

## Data Availability

All data produced in the present work are contained in the manuscript

## ACKNOWLEDGEMENTS

Not applicable.

## DATA SHARING

No additional data are available.

