## Supplementary File 2 for "Effectiveness and clinical relevance of kinesio taping in musculoskeletal disorders: A protocol for an overview of systematic reviews and evidence mapping"

**Supplementary File 2. Application of the GRADE framework in the systematic review without meta-analysis**

**Risk of bias**

The quality of each primary randomised controlled trials (RCTs) will be assessed using the Cochrane Risk of Bias assessment tool 2.0 (ROB 2.0).[1] The overall risk of bias will be judged as ‘low risk of bias’, ‘some concerns’ or ‘high risk of bias’. According to the GRADE guidelines, the summary of evidence will be downgraded if the RCTs is at a serious risk of bias.[2] The evaluation criteria are listed below:

(a) no serious risk of bias overall (0 downgrade): all RCTs are at low overall risk of bias.

(b) serious risk of bias overall (1 downgrade): at least one RCT is at some concerns for overall risk and the remaining RCTs are at low.

(c) very serious risk of bias overall (2 downgrade): at least one RCT is at high overall risk of bias and the remaining RCTs are at low or some concerns.

**Inconsistency**

The consistency of the summary of evidence will be considered by the percentage of RCTs that showed any same or similar enough differences (i.e., positive effect, no effect, and negative effect) out of all studies. A cut-off of 75% will be set for assessing the overall effect.[3] When there is only one outcome-related RCT included, the assessment will not be applicable due to substantial heterogeneity and lack of confidence. The evaluation criteria are listed below:

(a) no serious risk of inconsistency (0 downgrade): the percentage of RCTs that demonstrated the same effect is over 75%.

(b) serious risk of inconsistency (1 downgrade): the percentage of RCTs that demonstrated the same effect is less than or equal to 75%.

**Indirectness**

The indirectness of the summary of evidence may be due to differences in the measurement of comparisons, populations, interventions, comparators, and outcomes. Since we will manually check the inclusion of eligible comparisons, interventions, comparators and outcomes, the indirectness may exist in the remaining portion. The evaluation criteria are listed below:

(a) no serious risk of indirectness in populations (0 downgrade): the RCTs included participants with a diagnosis of musculoskeletal disorders only according to the International Classification of Diseases 11th revision (ICD-11).[4]

(b) serious risk of indirectness in populations (1 downgrade): the RCTs included participants with diagnoses of both musculoskeletal disorders and other disorders (e.g., diseases of the nervous, circulatory, genitourinary system) according to the ICD-11.[4]

**Imprecision**

The imprecision of the summary of evidence will be considered the associated 95% confidence interval (CI) (Wilson interval) of the percentage of RCTs out of all studies and the number of included participants.[3,5] The evaluation criteria are listed below:

(a) no serious risk of imprecision (0 downgrade): the lower limit of 95% CI of the percentage of RCTs is over 50% and the number of included participants is over 200.

(b) serious risk of imprecision (1 downgrade): the lower limit of 95% CI of the percentage of RCTs is less than or equal to 50% or the number of included participants is less than or equal to 200.

(c) very serious risk of imprecision (2 downgrade): the lower limit of 95% CI of the percentage of RCTs is less than or equal to 50% and the number of included participants is less than or equal to 200.

**Publication bias**

The publication bias of the summary of evidence will not be assessed since the funnel plots or other statistical methods is not applicable. However, in case actual evidence is discovered, we will consider downgrading according to the Cochrane Collaboration Handbook.[6]
