## Supplementary File 3 for "Effectiveness and clinical relevance of kinesio taping in musculoskeletal disorders: A protocol for an overview of systematic reviews and evidence mapping"

**Supplementary File 3. Calculation of clinical relevance**

The distribution-based methods for determining the minimal clinically important difference (MCID) will be used. Distribution-based approaches are defined based on statistical parameters reflecting statistical spread/variation and measurement accuracy of the outcome.[1-3] We will select the standard deviation (SD) to calculate the MCID and the formula is:

MCID=0.5*SD,

the SD represents the SD of pretreatment outcome scores of the control group of patient population studied. Considering the disadvantage that the calculation of the MCID is sample-dependent, the selection of SD should be rigorous.[1,4] Hence, the SD will be derived from the most representative randomised controlled trial (i.e., highest number of participants and lowest risk of bias) of the best systematic review (i.e., most comprehensive, most recent or up-to-date, highest methodological quality and lowest risk of bias, most complete reporting).
